# Pharmaceutical Product Recalls in Kenya (2016 to 2025): Trends in Quality Defects, Therapeutic Class Patterns, Manufacturing Gaps, and Regulatory Insights

**DOI:** 10.1101/2025.11.20.25340698

**Authors:** Alan Omondi odoyo

## Abstract

This retrospective study analyzes 171 pharmaceutical product recalls issued by the Pharmacy and Poisons Board (PPB) in Kenya between January 2016 and November 2025 to characterize patterns in medicine quality defects, therapeutic classes affected, manufacturing gaps, and regulatory implications. Recall notices were extracted from the PPB database and coded by year, therapeutic category, dosage form, product origin, and stated reason for recall. Of the 171 recalled products, 148 (86.5%) were due to quality-related defects, with physical and organoleptic abnormalities such as discoloration, spotting, molding, and tablet breakage representing 63 (36.8%) recalls, the largest single defect sub-category. Chemical and assay failures were observed in 34 (19.9%) recalls whereas 24 (14.0%) recalls were due to performance defects affecting dissolution, uniformity, and hardness. Anti-infectives particularly antibacterials and nervous-system products especially analgesics accounted for 52 (30.4%) and 37 (21.6%) of the recalled drug classes respectively, reflecting both their high market volume and vulnerability to quality test failures. Imported products accounted for 91 (53.0%) recalls, while locally manufactured products were 80 (47.0%). The observed upward trend in recall frequency is likely attributable to strengthened post-market surveillance rather than a true decline in product quality.

The findings highlight recurrent manufacturing and stability gaps and challenges, including inadequate in-process controls, insufficient supplier qualification, weak stability testing for WHO Climatic Zone IVb conditions, and suboptimal packaging and distribution practices. To address these gaps, the study recommends enhanced quality assurance frameworks enhanced by current GMP and quality management systems among manufacturers, expanded post-market surveillance and pharmacovigilance programs by regulators, strict enforcement of climate-appropriate stability and packaging requirements, and adoption of digital track-and-trace systems. Strengthened regulatory stringency, transparency and surveillance are essential to mitigating the circulation of substandard and falsified medicines and safeguarding public health in Kenya.

## 1. INTRODUCTION

Enforcement of pharmacovigilance and post-market surveillance (PMS) programs in addition to market authorizations and conducting current good manufacturing (cGMP) audits by drug regulatory agencies is essential in ensuring quality of pharmaceutical products available to consumers. There is limited industry-controlled production of pre-market data, rendering consumers unable to suspect quality deterioration of drugs, coupled with problematic drug prescription and consumption practices that exacerbate the already dire public health concern [1]. Recall of a marketed product is one of the most immediate corrective actions regulators and manufacturers can take when safety or quality defects are detected [2]. While pre-market surveillance is useful in the testing the quality of drugs, further subjection of drugs to different conditions in a controlled environment during stability studies does not fully cover the myriad of variables that these drugs are exposed to in the market. This is what makes post-market surveillance more important, albeit limited in its scope and intervention. In Kenya, the Pharmacy and Poisons Board (PPB) oversees regulatory control of human pharmaceuticals, including post-authorization monitoring, recalls and withdrawals [3]. According to PPB guidelines, recalls may be voluntary (initiated by the holder, often upon internal detection of a defect) or statutory (mandated by the Board after a defect is confirmed).

Drug recalls are essential to pharmacovigilance, helping to reduce risks from poor-quality medicines that could lead to adverse treatment outcomes. The PPB plays a vital role in communicating drug recall patterns through its website and social media platforms, which can reveal defective products and prompt companies to act more quickly in recall and withdrawal of affected batches [4]. The PPB uses a recall classification system similar to international standards: Class I–III recalls (based on urgency and risk) may be announced and are categorized as either voluntary or statutory. Class I recalls, for example, suggest serious health risks if not addressed, while Class II and III involve less severe defects. Generally, voluntary recalls often indicate internal quality issues identified by the manufacturer or through competitor reports, whereas statutory (Board-mandated) recalls follow PPB inspections or quality control laboratory tests that reveal noncompliance.

Globally, recall patterns in Kenya can be compared to those in similar low- and middle-income markets. For instance, Nigeria and Uganda have also experienced frequent recalls due to assay failures, contamination, and labeling errors, which indicate manufacturing issues [5] [6]. Strict adherence to WHO prequalification standards and Good Manufacturing Practices (GMP) for manufacturing, storing, and distributing pharmaceuticals results in lower defect rates, making drugs produced this way less likely to fail quality assurance tests. However, local manufacturers that produce generic drugs often have higher failure rates in tests, according to quality surveys [7]. In East Africa, harmonization efforts (EAC-MRH) are gradually raising standards, but recall data reveal that gaps still exist [8]. Therefore, reviewing Kenya’s pharmaceutical drugs’ recall and withdrawal history over the last decade offers valuable insight into both national and regional medicine quality challenges.

## 2. METHODS

We conducted a retrospective, systematic analysis of drug recall data provided by the PPB, covering the period between 2016 and November 2025. Each entry contained details on the product name, batch number, manufacturer, reason for recall and status of recall/withdrawal. The PPB website publishes annual recall lists summarizing each product recalled (including name, formulation, batch number, manufacturer, and reason). Each recall entry was extracted from the PPB website and compiled for analysis. The key variables for each recall were year of recall, product category, dosage form, nature of defect (e.g., assay out-of-specification, dissolution failure, microbial/particulate contamination, color change or physical defect, labeling error, packaging defect, stability failure, etc.), and product origin (local manufacturer vs. imported). Recall causes were collated verbatim from the notices and then grouped into broader categories for analysis. The data was compiled and analyzed in Microsoft Excel 2024 worksheets. Data was presented in tables and charts. A potential limitation is reliance on the data available on the PPB website, which may not encompass all the recalls or problematic drugs in the market.

## 3. RESULTS AND ANALYSIS

### 3.0 Overview of Recall Characteristics

There were a total of 171 product recalls in the period studied, from January 2016 to November 2025. Out of these, 80 recall cases representing 47.0% were locally manufactured products, while 91 cases (53%) were imported products. Recall trend indicated a steady rise in the number of recalls, dipping from 2020-2022 and rising again. This points to a possible increase in post-market analysis and increased pharmacovigilance programs characterized by greater reporting of quality defects. There were very few recalls in the early years (2016-2018) as opposed to the later years (2023-2025), which corresponds with reports on increased market volume, increased sampling and testing of drugs by quality control laboratories, and growing GMP compliance pressures on manufacturers. Three main categories of recall trends are analyzed. Understanding these categories provides insight into the recall characteristics of the drugs, which may inform policy formulation. The categories include distribution by category, looking at the dominant classes of drugs recalled, reasons for recall, and distribution by country of origin. Overall, the dataset shows that quality-related defects accounted for the overwhelming majority (86.5%) of all recall reasons, followed by GMP/manufacturing failures and labelling errors (7.6% each). Only seven products (4.1%) were recalled due to pharmacovigilance or clinical safety concerns, and four (2.3%) were withdrawn for administrative or voluntary reasons.

### 3.1 Distribution by Therapeutic Category

Table 1 below shows the distribution category of drugs recalled by the PPB 2016-2025 by body system/ drug class.

**Table 1.** Body System and Drug Class Distribution.

|  | Body System/ Drug class | Number of samples n | Percentage (%) |
| --- | --- | --- | --- |
| **1** | Anti-infectives | 52 | 30.4 |
|  | a. Antibacterials | 27 | 15.8 |
|  | b. Anthelmintics | 10 | 5.8 |
|  | c. Antivirals | 4 | 2.3 |
|  | d. Antifungals | 4 | 2.3 |
|  | e. Antimalarials | 1 | 0.6 |
|  | f. Antimycobacterials | 1 | 0.6 |
|  | g. Mixed anti-infectives | 5 | 2.9 |
| **2** | Nervous system | 37 | 21.6 |
|  | a. Analgesics | 21 | 12.3 |
|  | b. Psychotropics | 7 | 4.1 |
|  | c. Anesthetics | 2 | 1.2 |
|  | d. Anti-epileptics | 2 | 1.2 |
|  | e. Anti-inflammatory drugs | 5 | 2.9 |
| **3** | Cardiovascular system | 16 | 9.4 |
|  | a. Antihypertensives | 14 | 8.2 |
|  | b. Antithrombotics | 2 | 1.2 |
| **4** | Nutritional products | 13 | 7.6 |
|  | a. Minerals & Vitamins | 4 | 2.3 |
|  | b. Vitamins | 2 | 1.2 |
|  | c. Electrolytes | 3 | 1.8 |
|  | d. Minerals | 1 | 0.6 |
|  | e. Nutrient mixtures | 1 | 0.6 |
|  | f. Waters | 1 | 0.6 |
| **5** | Genitourinary system | 11 | 6.4 |
|  | a. Contraceptives | 9 | 5.3 |
|  | b. Sexual dysfunction drugs | 1 | 0.6 |
|  | c. Uterotonics | 1 | 0.6 |
| **6** | Gastrointestinal system | 11 | 6.4 |
|  | a. Antiulcer drugs | 8 | 4.7 |
|  | b. Laxatives | 1 | 0.6 |
|  | c. Spasmolytics | 1 | 0.6 |
|  | d. Antiemetics | 1 | 0.6 |
| **7** | Anticancer agents | 1 | 0.6 |
|  | a. Anticancer agents | 1 | 0.6 |
| **8** | Respiratory system | 8 | 4.7 |
|  | a. Respiratory system | 8 | 4.7 |
| **9** | Skin preparations | 6 | 3.5 |
|  | a. Skin preparations | 6 | 3.5 |
| **10** | Musculoskeletal system | 2 | 1.2 |
|  | a. Muscle relaxants | 2 | 1.2 |
| **11** | Miscellaneous | 14 | 8.2 |
|  | a. Antiseptics/disinfectants | 6 | 3.5 |
|  | b. Medical devices/equipment | 6 | 3.5 |
|  | c. Vaccines | 1 | 0.6 |
|  | d. Immunomodulatory agents | 1 | 0.6 |
|  | Total | 171 | 100 |

As represented in Table 1, certain therapeutic categories are disproportionately affected by defective products. The dominant classes of drugs affected by recalls are anti-infectives and the nervous system drug categories. Out of the 171 recalls during the period of study, 52 (30.4%) of recalls were anti-infectives, with anti-bacterials alone accounting for 27 (15.8%) products. Such high recall rates indicate are definitely a public health concern considering these classes of drugs are prone to resistance development. The second-highest category of drugs recalled was nervous system drugs, particularly analgesics. More than a fifth of the recalled drugs (21.6%) were nervous system drugs, with 21 (12.3%) being analgesics. This could be attributed to the heavy presence of common over-the-counter (OTC) pain relievers, many of which are locally manufactured. Recalls for cardiovascular drugs were 9.4% of total recalls, while nutritional supplements accounted for 7.6% of recalls. A significant public health concern is posed by the recalls of genitourinary products, especially contraceptives, which accounted for 6.4% of the total recalls, posing a risk of potential unwanted pregnancies and spread of STIs. Anticancer agents and uterotonics had the lowest frequency of recalls (0.6% each).

Looking at the recall by therapeutic category, insights that can be drawn from the data reveal that recalls for these products are not random, but a reflection of systemic patterns. The most frequent recalls are clustered around drugs that are widely used and required in high volumes. These drugs also happen to have a large representation of generic therapeutic drugs manufactured to meet the high demand. Medicines requiring complex manufacturing processes such as anti-cancers were recalled less frequently, but when they do, they often indicate more serious systemic GMP gaps.

### 3.2 Reasons for Recall

The PPB website provides a reason for each recall. Table 2 below categorizes recalls by reasons.

**Table 2.** Distribution of Recall Reasons by Category (n=171)

|  | Reason for recall | Sub-category | Count n |
| --- | --- | --- | --- |
| **1** | Quality Defects | 1a. Physical/ Organoleptic (color change, spots, molding, broken tablets, appearance) | 63 |
|  |  | 1b. Chemical / Assay / Related Substances OOS | 34 |
|  |  | 1c. Performance / Dissolution / Hardness / Weight / Uniformity/ Weight Variation | 24 |
|  |  | 1d. Microbiological / Molds / TAMC / Contamination | 4 |
|  |  | 1e. Stability / OOT / Degradation / GTI | 9 |
|  |  | 1f. Contamination / Foreign Particles / Precipitates | 14 |
|  |  | Total (Quality Defects) | 148 |
| **2** | GMP / Manufacturing Failures | 2. Process Defects / Mix-ups | 13 |
| **3** | Labeling & Packaging Errors | 3. Missing / Incorrect / Mislabeled | 13 |
| **4** | Safety / Pharmacovigilance Issues | 4. Adverse Event / Therapeutic / Clinical Risk | 7 |
| **5** | Administrative / Regulatory | 5. Voluntary / Market Authority Recall | 4 |
|  | **Total** |  | **171** |

Data was sorted and classified according to the reason for recall. The reasons for recall were either quality defects, GMP/manufacturing failures, labelling/packaging errors, safety/pharmacovigilance issues, or administrative/regulatory reasons. The reasons for recall provide direct insights into the issues with drugs marketed in the country.

#### Quality Defects

The largest category of reasons for recall was quality defects, accounting for 148 (86.5%) recalls. Out of this, the largest sub-category of recalls was due to physical/organoleptic defect, accounting for 36.8% of the total recalls over the 10 years. Examples of these include color change, spots, molding, broken tablets, and appearance. The quality defect reasons were summarized and grouped into the sub-categories 1A to 1F as shown in Table 3 below.

**Table 3.** Quality Defects Sub-classifications.

| **1A. Physical / Organoleptic Defects (n = 63)** |  |  |  |
| --- | --- | --- | --- |
| **Subcategory 1a** | **Description/Examples** | **Count (n)** | **%** |
| 1a-i. Color Change / Discoloration | Tablets/capsules/powder changing color, fading, darkening, yellowing, pink → brown, blackening | 25 | 39.7 |
| 1a-ii. Spots / Black Spots / Brown Dots / Mottling | Visible dark or colored spots, specks, brown dots, black spots, mottled appearance | 13 | 20.6 |
| 1a-iii. Moulding / Mold-like Defects | Tablets or suspensions showing molds, molding, capping/lamination issues | 9 | 14.3 |
| 1a-iv. Broken / Cracked / Chipped Tablets / Capsules | Tablets crumbling, broken, cracked along score lines, or stuck to blister | 3 | 4.8 |
| 1a-v. Appearance / Organoleptic Defects (others) | Lump formation, unpleasant odor, off-white / off-color suspensions, caking, clumping | 5 | 7.9 |
| 1a-vi. Hard to Break / Coating Loss / Clumping Issues | Tablets hard to break, clumping of coating, loss of coat color | 3 | 4.8 |
| 1a-vii. Miscellaneous / Combination Defects | Multiple overlapping appearance issues reported in one batch | 5 | 7.9 |
| **Total** |  | **63** | **100** |
| **1B. Chemical / Assay / Related Substances OOS (n = 34)** |  |  |  |
| **Sub-Category 1b** | **Description / Examples** | **Count (n)** | **%** |
| 1b-i. Assay Failures / API Content OOS | Active pharmaceutical ingredient outside specification limits (too high / too low) | 24 | 70.6 |
| 1b-ii. Related Substances / Impurity Failures | Out-of-specification for degradation products, unknown impurities, GTIs | 8 | 23.5 |
| 1b-iii. pH / Other Chemical Parameters | pH outside specification, unusual chemical changes | 2 | 5.9 |
| **Total** |  | **34** | **100** |
| **1C. Performance / Dissolution / Hardness / Uniformity / Weight Variation (n = 24)** |  |  |  |
| **Sub-Category** | **Description / Examples** | **Count (n)** | **%** |
| 1c-i. Dissolution Test Failures | Tablets/capsules failing dissolution specifications | 14 | 58.3 |
| 1c-ii. Assay-related Performance Issues | Combination of assay + performance deviations (affecting dissolution, content uniformity) | 4 | 16.7 |
| 1c-iii. Uniformity / Weight Variation Failures | Weight variation or content uniformity outside spec | 8 | 33.3 |
| 1c-iv. Hardness / Thickness / Friability Failures | Tablets failing hardness, friability, thickness tests | 6 | 25 |
| **Total** |  | **32** | **100** |
| **1D. Microbiological / Mold / TAMC / Contamination (n = 4)** |  |  |  |
| **Sub-Category** | **Description / Examples** | **Count (n)** | **%** |
| 1d-i. Mold / Fungal Contamination | Visible molds, mold-like spots, microbial growth in product | 3 | 75 |
| 1d-ii. Microbial Count / TAMC / Total Yeast & Mold Count | Out-of-specification microbial limits in finished product or raw materials | 1 | 25 |
| **Total** |  | **4** | **100** |

| **1E. Stability Failures / OOT / Degradation / GTIs (n = 9)** |  |  |  |
| --- | --- | --- | --- |
| **Sub-Category** | **Description / Examples** | **Count (n)** | **%** |
| 1e-i. Out-of-Trend / OOT Results | Stability results trending outside expected limits (assay, dissolution, related substances) | 3 | 33.3 |
| 1e-ii. Degradation / Impurity Formation | Degradation of API or formation of impurities over time in stability study | 5 | 55.6 |
| 1e-iii. General Stability Failures | Other stability-related deviations not classifiable above (content, potency, appearance changes in stability) | 1 | 11.1 |
| **Total** |  | **9** | **100** |
| **1F. Contamination / Foreign Particles / Precipitates (n = 14)** |  |  |  |
| **Sub-Category** | **Description / Examples** | **Count (n)** | **%** |
| 1f-i. Visible Particles / Precipitates in Solution | Foreign particles or precipitates seen in suspensions, powders, or reconstituted solutions | 5 | 35.7 |
| 1f-ii. Contaminated / Mixed-up Packs | Cross-contamination or mix-up with other products/blisters | 4 | 28.6 |
| 1f-iii. Packaging / Component-related Contamination | Contamination due to defect in primary/secondary packaging or port defect | 5 | 35.7 |
| **Total** |  | **14** | **100** |

Color changes/discoloration accounted for 25 (39.7%) recalls making it the single most common visible defect, pointing to issues in suboptimal packaging, stability, inadequate environmental controls, or excipient incompatibility. Recalls were due to presence of black dots, spots and mottling indicating issues related to contamination, poor mixing, granulation defects, or the use of substandard materials were 13 (20.6%). Other physical defects included molding and fungal growth and broken, chipped, or cracked tablets indicating moisture ingress or inadequate preservative systems and poor mechanical strength and compression failures respectively. The physical/organoleptic defects point to poor environmental control, inadequate stability testing, and substandard packaging, especially in high volume generic drugs.

Chemical/assay/impurity failures led to a fifth of all recalls during the study period. Out of this, 24 (70.6%) were due to assay-related failures. Key trends indicate API content being outside specification as the major contributor to these substandard medicine, reflecting deficiencies in raw material quality and in-process controls. Additionally, impurity and degradation failures, accounting for 8 (23.5%) of the defects in this category, reinforce concerns about stability, especially in tropical climatic conditions. These chemical defects strongly signal GMP lapses, instability, and manufacturing process inconsistencies. They have direct therapeutic implications—products with sub-potent active pharmaceutical ingredients may lead to treatment failure and resistance development, especially observed in antibiotics.

Recalls due to performance, dissolution, hardness and uniformity failures were 24 (14.0%) of all recalls. Out of these, about 14 (58.3%) were due to dissolution failures. This means that the drugs had formulation weaknesses, such as poor-quality binders/disintegrants, inadequate granulation, or poor compression control. Such defects are more prevalent in manufacturers with weak quality-by-design (QbD) frameworks. Issues with microbiological contamination were few (2.3% of the cases), while stability-related failures led to 9 (5.3%) recalls. These issues are critical because microbial contamination poses direct safety risks, while stability issues suggest insufficient accelerated and long-term stability studies, or inappropriate packaging for Kenya’s hot and humid climate (Climatic Zone IVb). Other reasons for recall included foreign particles, precipitates, and packaging-linked contamination (8.2%), GMP/manufacturing process failures (7.6%), labelling and packaging errors (7.6%), safety and pharmacovigilance-triggered recalls (4.1%), and administrative and regulatory recalls (2.3%).

The patterns from the drug recalls by the PPB provide valuable insights into the country’s pharmaceutical industry that need to be addressed. The first issue is the high prevalence of physical and chemical defects. Together accounting for nearly two-thirds of all recalls, these defects clearly signal stability weaknesses, manufacturing inconsistencies, substandard packaging materials, and deficient quality control systems. Second, there is a notable vulnerability of generic, high-volume, locally produced medicines observed from the heavy presence of anti-infectives and analgesics, mostly generics. This indicates high production volumes, price-driven manufacturing shortcuts, and limited investment in advanced QC technologies.

The rise in recalls from 2016–2025 is a positive regulatory indicator, reflecting stronger PPB risk-based sampling, improved post-market surveillance, and better reporting channels. Finally, defective antibiotics, contraceptives, cardiovascular drugs, and pediatric formulations represent the most critical risks due to treatment failure, antimicrobial resistance, reproductive health impact, and toxicity potential. In summary, the recall data from 2016–2025 clearly demonstrate that most defective products in Kenya fail not because of acute safety issues, but due to preventable quality defects—primarily physical, chemical, and performance failures. These defects are characteristic of systemic lapses in cGMP adherence, weak stability testing programs, and insufficient quality evaluation programs. Anti-infectives and analgesics are disproportionately affected, raising significant public health concerns. The trends strongly support the need for strengthening GMP enforcement, stability testing under Climatic Zone IVb, packaging and formulation robustness, post-market surveillance and sample testing, industry-wide quality-by-design (QbD) adoption, and digital recall and defect-trend analytics for assured quality, safety and efficacy of human pharmaceuticals.

## 4. DISCUSSION

The analysis of 171 pharmaceutical product recalls issued by the Pharmacy and Poisons Board (PPB) between 2016 and 2025 reveals systemic quality challenges within Kenya’s pharmaceutical product market. The dominant presence of physical, chemical, and performance-related defects, which collectively account for over 80% of all recalls, demonstrates that most product withdrawals arise from preventable manufacturing deficiencies and failures. This pattern is consistent with research in other low- and middle-income countries (LMICs), where weak GMP adherence and unstable supply chains contribute significantly to the circulation of substandard medicines [9] [10].

### 4.1 Quality Defects and Manufacturing Weaknesses

Physical and organoleptic defects (e.g., discoloration, molding, spots on tablets) comprised the largest defect category. These failures often reflect gaps in stability studies, moisture control, and packaging suitability, especially in Climatic Zone IVb conditions, which require robust stability data at 30°C/75% RH [11]. The high rate of discoloration and impurity formation further suggests probable chemical reactions (drug-excipient degradation, drug-excipient interaction or drug-impurity interaction), microbiological contamination with yeast, mold or bacteria, and physical contamination either from dust, dirt, hair, metal chippings or foreign material.

Chemical and assay failures, representing approximately 20% of the recalls, indicate significant lapses in in-process controls, raw material quality assurance, and batch uniformity. SFMs pose major therapeutic risks, particularly in antimicrobials prone to development of resistance. Studies in East Africa have repeatedly shown assay failures in commonly used antibiotics, contributing to antimicrobial resistance (AMR) due to under-dosing and treatment failure [12] [13]. These findings align with WHO’s broader concern that substandard medicines play a substantial role in global AMR escalation [7]. Performance-related failures such as dissolution and uniformity deviations point towards weak formulation processes, improper granulation, or poor compression control. These issues are frequently observed in generic manufacturing environments where cost pressures lead to inadequate investment in quality-by-design (QbD) and process analytical technologies (PAT) [1]. The clustering of these defects among analgesics and antibiotics suggests systemic weaknesses within high-volume local manufacturing lines.

### 4.2 Therapeutic Categories and Public Health Implications

The disproportionate recall of anti-infectives and analgesics underscores the vulnerability of high-demand therapeutic classes that dominate Kenya’s pharmaceutical market. Similar recall patterns have been documented in Asia and West Africa, where high-volume generics tend to experience higher defect rates due to pressure to reduce manufacturing costs [4] [6]. The recall of contraceptives and cardiovascular medications is particularly concerning because failures in these categories have profound public health consequences, including unintended pregnancies, therapy interruptions, and preventable morbidity.

The steady increase in recall numbers over the study period likely reflects improved regulatory oversight. Kenya’s PPB has strengthened post-market surveillance, ramped up risk-based sampling, and leveraged laboratory testing capacity in line with WHO Global Surveillance and Monitoring System (GSMS) recommendations. Similar upward recall trends have been observed in South Africa, India, and Ghana following regulatory reforms [14] [15]. Therefore, the increase in recalls may represent improved detection, not necessarily worsening product quality.

Globally, the most common reasons for drug recalls including assay failures, contamination, dissolution issues, and labeling errors mirror the Kenyan findings [16] [17]. However, the frequency of physical defects and stability-linked failures is notably higher in Kenya, consistent with climatic, infrastructural, and cGMP adherence challenges typical of LMIC pharmaceutical sectors [10] [19]. This contextual insight highlights the need for climate-adapted formulations, stronger environmental control systems, and improved packaging standards. Taken together, the recall data suggest that Kenya’s drug quality issues are largely manufacturing-driven, reflecting insufficient GMP maturity across segments of the pharmaceutical industry, inadequate stability testing, weak supplier qualification and raw material verification, limited adoption of QbD and process validation frameworks, and a growing but still insufficient regulatory surveillance. These findings provide a strong evidence base for policy intervention. Accurate prediction and prevention of pharmaceutical product defects is attainable through multipronged approaches including institutionalization of quality manufacturing systems broadly classified into process performance and product quality monitoring system, corrective action and preventive action (CAPA) system, change management system and management reviews [18].

## 5. CONCLUSION

This study demonstrates that drug recalls in Kenya between 2016 and 2025 predominantly stem from quality and manufacturing defects, particularly among high-volume generic medicines. Deficiencies in physical appearance, assay/content uniformity, stability, and performance represented the bulk of the recalls, indicating systemic GMP shortcomings and insufficient investment in robust formulation and quality manufacturing systems. Although recall frequency has increased over the study period, this likely reflects improvement in PPB surveillance capability and regulatory enforcement rather than a direct deterioration in product quality. The findings underscore the need for enhanced GMP compliance, strengthened stability programs suited for hot and humid climates, better raw material controls, increased regulatory capacity for proactive post-market surveillance and strengthened pharmacovigilance programs so as to collectively ensure quality medicines reach the population [19]. By addressing these gaps, Kenya can significantly reduce the prevalence of SFMs and enhance patient safety in line with WHO global quality assurance standards.

## 6. RECOMMENDATIONS

From this study, several recommendations are made. First, there is need to strengthen cGMP compliance and regulatory enforcement through increased frequency of GMP audits with risk-based prioritization, mandating corrective and preventive action (CAPA) verification audits for repeat offenders and strengthening penalties for non-compliance to deter negligent manufacturing practices. Stability testing under climatic zone IVb testing requirements should also be improved. This can be achieved through requiring manufacturers to generate region-specific stability data, promoting the use of packaging that provides improved moisture and light protection, and implementing periodic stability monitoring for products already on the market.

Manufacturing quality systems need to be enhances by implementing stricter raw material supplier qualification protocols and encouraging adoption of Quality-by-Design (QbD) and Process Analytical Technology (PAT) for consistent batch quality. Implementing stricter raw material supplier qualification protocols and training local manufacturers on advanced formulation, granulation, and compression technologies can also improve the overall manufacturing quality systems to reduce quality defects. While there is improved post-market surveillance as indicated by the increasing number of recalls, the PMS and field testing needs to be strengthened by increasing random sampling of high-risk therapeutic categories such as anti-infectives and contraceptives., implementing mobile field-testing technology (e.g., Minilab, TLC), as recommended by WHO, and developing a national digital PMS dashboard for real-time defect tracking.

The problem with mislabeling can be addressed through improving labelling and packaging controls. Suggestions include enforcing strict version control of artworks and packaging components, adopt automated vision inspection systems to reduce human error, and standardize packaging requirements for products prone to degradation. The government can also establish a national recall analytics and early warning system using the recall trend data to generate predictive indicators for defect recurrence, share industry-wide anonymized quality data to enhance transparency and prevention, and implement manufacturer-level periodic quality performance scoring. Finally, pharmacists and other consumer healthcare workers should be educated on recognizing early signs of product defects while strengthening public awareness campaigns about reporting suspected substandard medicines. These measures will not only protect consumers from harmful products but also contribute to the realization of the government’s Universal Healthcare vision.

## Data Availability

All data produced are available online at https://web.pharmacyboardkenya.org/recalls-and-withdrawals/

